# Volumetric quantification of wound healing by machine learning and optical coherence tomography in adults with type 2 diabetes: the GC-SHEALD RCT

**DOI:** 10.1101/2021.06.30.21259754

**Authors:** Yinhai Wang, Ramzi Ajjan, Adrian Freeman, Paul Stewart, Francesco Del Galdo, Ana Tiganescu

## Abstract

Type 2 diabetes mellitus is associated with impaired wound healing, which contributes substantially to patient morbidity and mortality. Glucocorticoid (stress hormone) excess is also known to delay wound repair. Optical coherence tomography (OCT) is an emerging tool for monitoring healing by “virtual biopsy”, but largely requires manual analysis, which is labour-intensive and restricts data volume processing. This limits the capability of OCT in clinical research.

Using OCT data from the GC-SHEALD trial, we developed a novel machine learning algorithm for automated volumetric quantification of discrete morphological elements of wound healing (by 3mm punch biopsy) in patients with type 2 diabetes. This was able to differentiate between early / late granulation tissue, neo-epidermis and clot structural features and quantify their volumetric transition between day 2 and day 7 wounds. Using OCT, we were able to visualize differences in wound re-epithelialisation and re-modelling otherwise indistinguishable by gross wound morphology between these time points. Automated quantification of maximal early granulation tissue showed a strong correlation with corresponding (manual) GC-SHEALD data. Further, % re-epithelialisation was improved in patients treated with oral AZD4017, an inhibitor of systemic glucocorticoid-activating 11β-hydroxysteroid dehydrogenase type 1 enzyme action, with a similar trend in neo-epidermis volume.

Through the combination of machine learning and OCT, we have developed a highly sensitive and reproducible method of automated volumetric quantification of wound healing. This novel approach could be further developed as a future clinical tool for the assessment of wound healing e.g. diabetic foot ulcers and pressure ulcers.

**Disclosure Summary:** I certify that neither I nor my co-authors have a conflict of interest as described above that is relevant to the subject matter or materials included in this Work.

## Introduction

In 2017/2018, the UK National Health Service (NHS) treated 3.8 million people with wounds, 1 in 3 of which failed to heal within a year (*1*). Direct costs were over £8 billion, with the majority for chronic wound care. Of concern, annual wound prevalence has increased by over 70% between 2012 and 2018, highlighting the urgent need to improve monitoring, management and treatment (*1*).

The financial burden of chronic wounds is mainly attributed to amputations in patients with diabetes (*2*). Since 1980, global rates of type 2 diabetes mellitus (T2DM) have more than quadrupled and the majority of countries remain unlikely to attain United Nation targets (*3, 4*). Chronic wounds (e.g. diabetic foot ulcers) affect around 5% of people with diabetes at any one time and have a 5-year mortality of up to 55% (*5, 6*). Affected individuals have a poor quality of life (*2, 7*) and the indirect costs associated with lost productivity are much greater than those of treatment (*8-10*).

Stress hormone (glucocorticoid) excess also leads to chronic wounds (*11, 12*). Glucocorticoids affect virtually every phase of wound healing, including inflammation (*13, 14*), re-epithelialization (*15, 16*), granulation tissue formation (*16)* and extracellular matrix (collagen) synthesis, processing and remodelling (*17, 18*). Local glucocorticoid levels are regulated by 11β-hydroxysteroid dehydrogenase (11β-HSD) isozymes which activate (11β-HSD1) and deactivate (11β-HSD2) both natural and synthetic corticosteroid forms (*19-23*). Through these enzymes, peripheral exposure to glucocorticoids is maintained independently of circulating hormone levels (*24*). We previously found increased 11β-HSD1 during normal mouse wound healing (*20*) and Brazel *et al*. recently found this to be exacerbated in diabetic (*db*/*db*) mice (*25*). Subsequently we (and others) reported accelerated healing in aged 11β-HSD1 knockout mice and following topical 11β-HSD1 inhibition in healthy mice and those treated with the oral glucocorticoid corticosterone (*21, 22, 26*). Improved wound healing following 11β-HSD1 blockade has also been found in animal models of diabetes (*27*) but this had not been explored in man.

Our recent double-blind, randomised controlled trial (GC-SHEALD) in adults with T2DM treated with the selective 11β-HSD1 inhibitor AZD4017 for 35 days found that wounds were 48% smaller at day 2 compared to placebo (*28*). This was based on maximal early granulation tissue width by optical coherence tomography (OCT), although no difference was found in maximal blood clot / scab depth at day 7 post-wounding (early granulation tissue was undetectable in day 7 wounds).

Optical coherence tomography (OCT) is a real-time tomographic imaging technique using low intensity infrared light to visualise living tissues. This non-invasive method enables high-resolution two-and three-dimensional cross-sectional imaging (analogous to histology) and is a validated method of monitoring skin structure and wound healing (*29-33*). However, OCT application has so far been restricted to manual annotation of two-dimensional scans which is time consuming, restricts the number of samples or time points that can be processed and limits scientific insights.

Here, we report the novel application of machine learning to identify and quantify key morphological features of wound healing in 3D, validated by manual two-dimensional analysis. This was validated using OCT outputs from the GC-SHEALD pilot phase 2b trial to analyse the wound healing response to systemic 11β-HSD1 inhibition in patients with T2DM.

## Results

### Machine Learning

#### Reliability

##### Wound Morphology

An illustrative example of typical machine-annotated outputs from days 2 and 7 post-wounding are shown (Figure 1). Wound morphology was classified into 4 sub-types based on key morphological features; 1) early granulation tissue, exhibiting a “honeycomb” appearance as fibrinolysis breaks down the clot and is replaced with early extracellular matrix, 2) late granulation tissue, displaying a uniform horizontal alignment, 3) neo-epidermis, extending from the wound margins at day 2 and adjoining beneath the clot by day 7 and 4) clot, forming a plug over the wound.

**Fig. 1.**
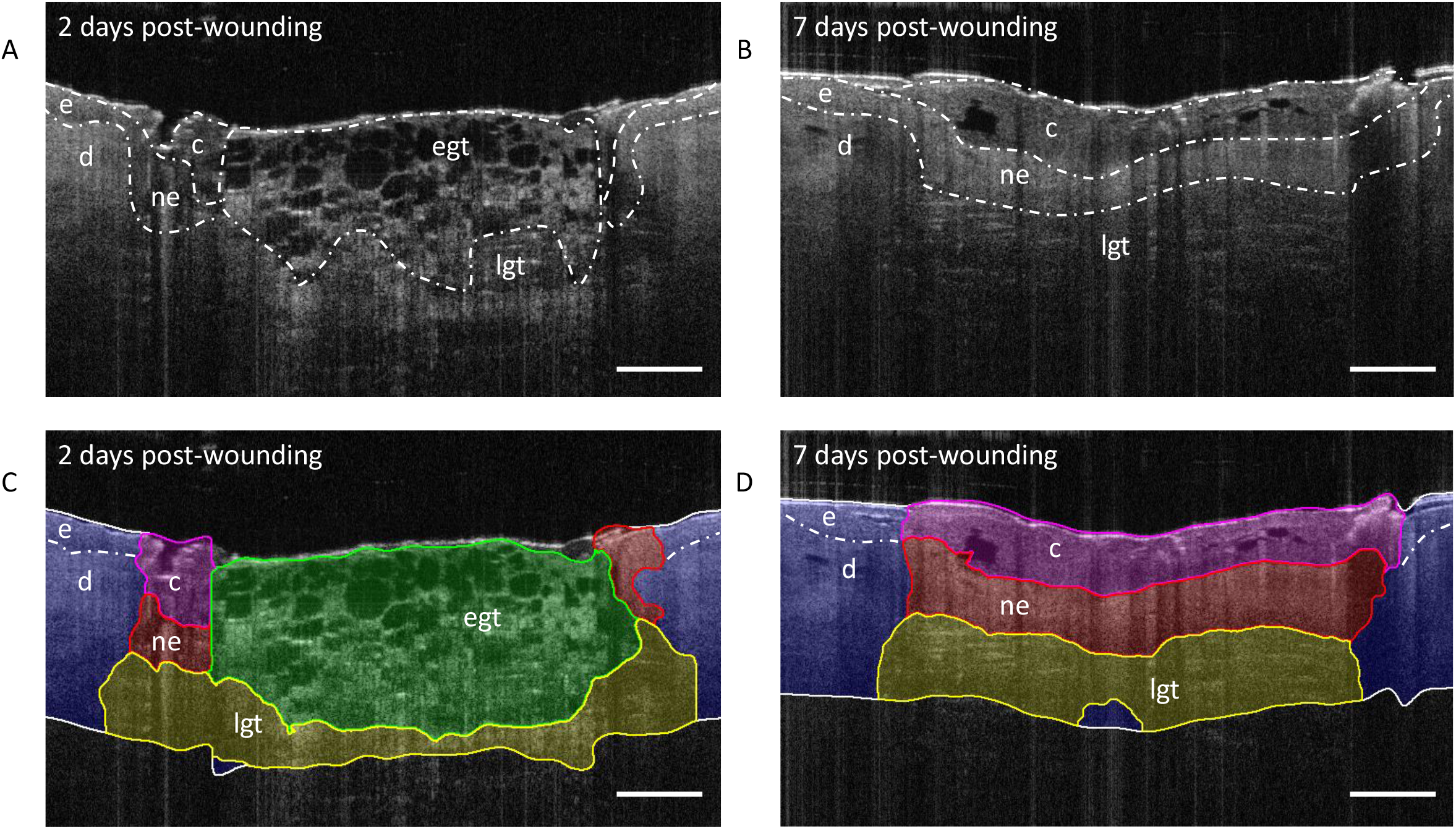
Machine learning outputs in day 2 and 7 wounds. Representative original OCT scan outputs (A, B) and with automated annotation following machine learning (C, D). c = clot, d = dermis (intact tissue), e = epidermis, get = early granulation tissue, lgt = late granulation tissue, ne = neo-epidermis. Scale bar = 500µm.

Automated annotation reliability was assessed for all scans. Investigator agreement with machine annotation accuracy (inter-observer reliability) was excellent (ICC>0.8) for all sub-types with ICC>0.95 for early granulation, late granulation and neo-epidermal tissue and ICC>0.85 for clot tissue. Altman plots, bias and limit of agreement for reliability of wound sub-type annotations are shown in Figure 2.

**Fig. 2.**
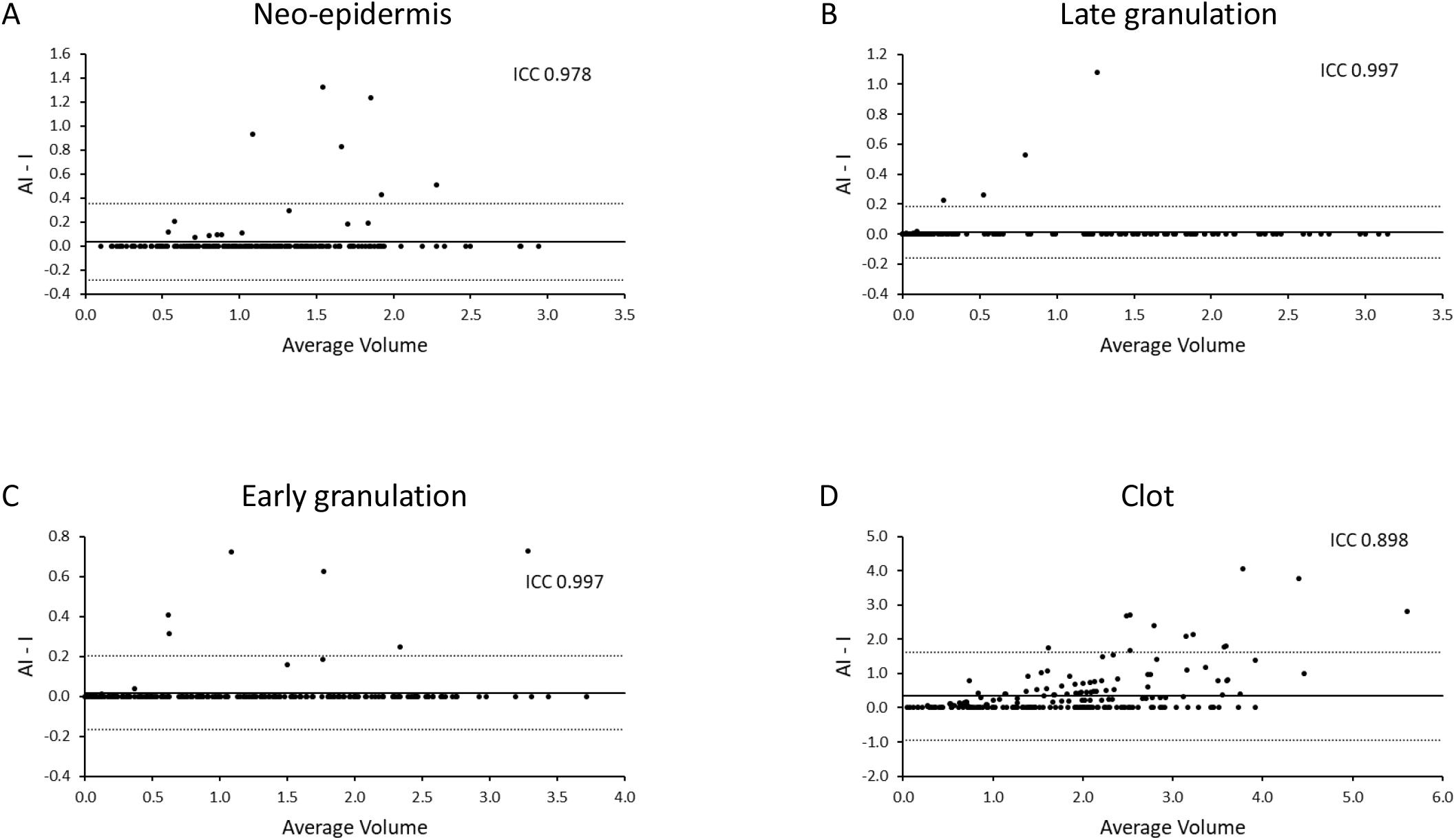
Machine learning reliability. Altman plots neo-epidermis (A), late granulation tissue (B), early granulation tissue (C) and clot (D) are shown. Intraclass correlation coefficients (ICC) and 95% limits of agreement (LoA) are displayed on each plot. Solid lines indicate the bias values and dotted lines (+2SD and −2SD) the LoA. N = 204.

##### Validation

A strong correlation (r=0.81, p<0.001) was observed between machine learning outputs and manual measurement of maximal early granulation tissue width (mm) at treatment days 2 and 30 (Figure 3a). However, this was not observed between machine learning outputs and manual measurement of maximal clot depth at treatment days 7 and 35 (Figure 3b).

**Fig. 3.**
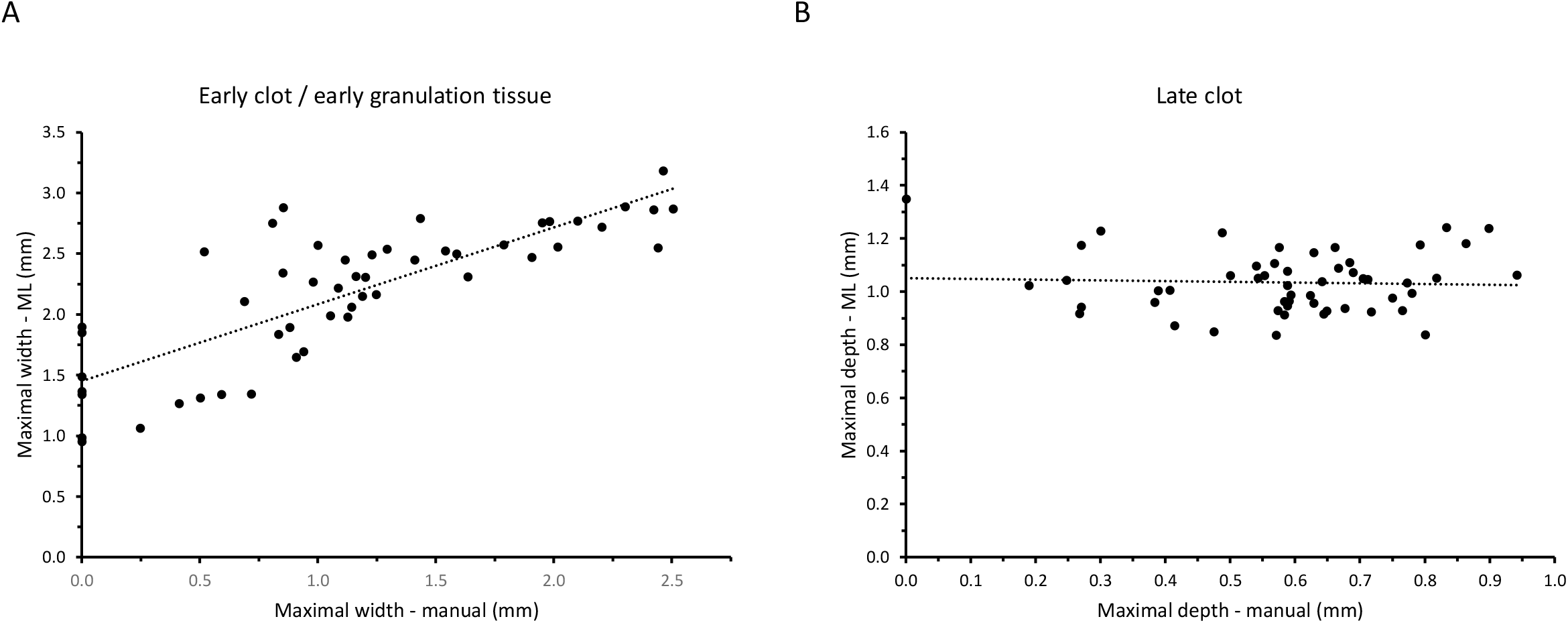
Machine learning validation. Correlations between machine learning outputs for maximal early granulation tissue (A, N = 51) and maximal clot depth (B, N=50) with manual measurement.

### Wound Morphology

#### Gross Morphology and Re-epithelialization

Gross wound area was largely comparable between treatments and time points. Using this indicator, healing improved from 37% to 49% between day 2 and day 7 in the placebo (PBO) group (p<0.05) in biopsies conducted at day 0 (set 1), but no improvement was observed between day 2 and day 7 wounds in the second round of biopsies conducted on treatment day 28 (set 2). Similarly, no treatment effect was discernible at either time point (Figure 4a and 4c).

**Fig. 4.**
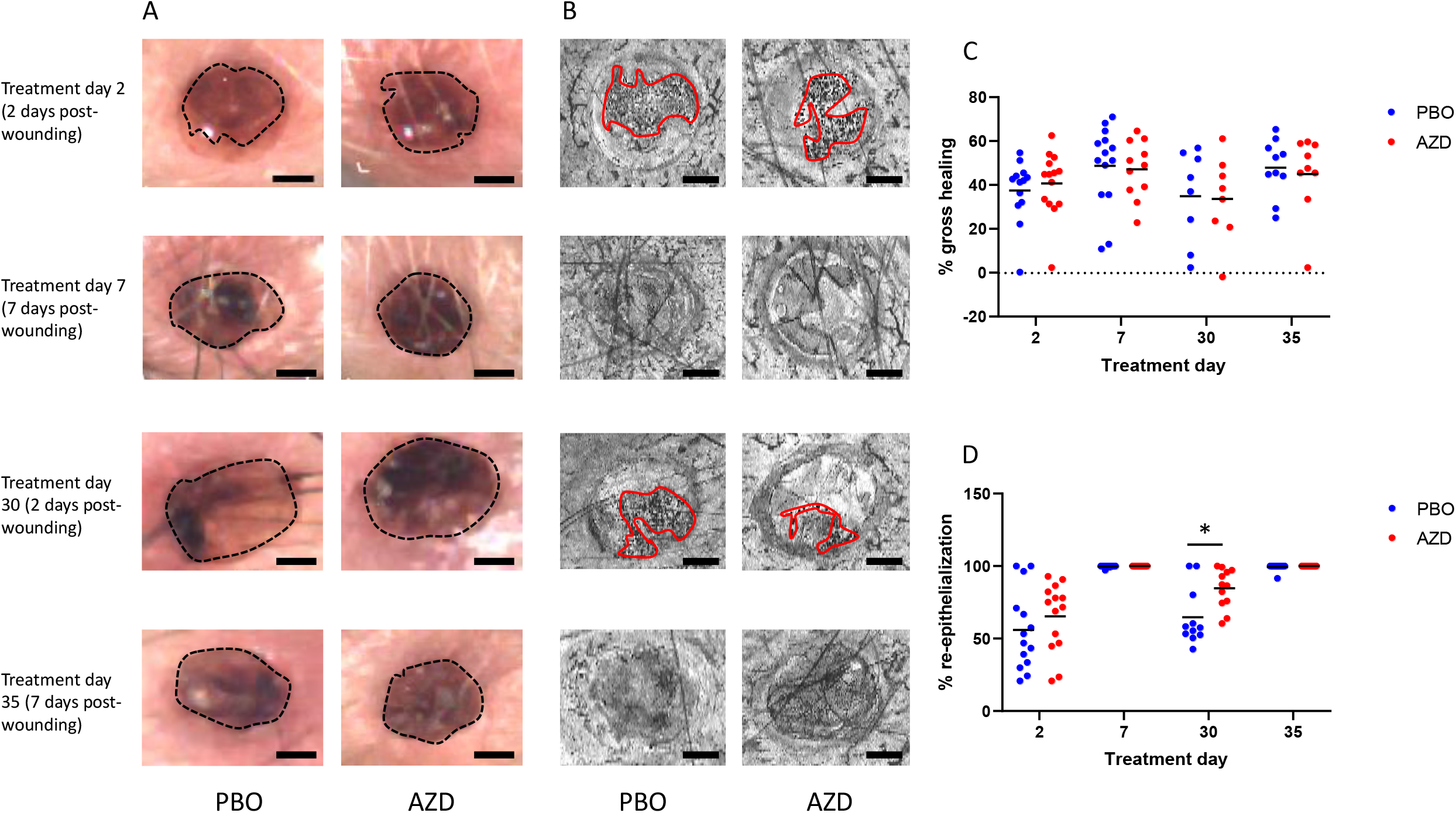
Gross Morphology and Re-epithelialization. Representative images for gross morphology (A) and re-epithelialisation by manual OCT (B), quantified manually as % gross healing (C) and % re-epithelialisation (D).

Manual analysis of matched OCT images revealed a large increase in % re-epithelialization (mean±SD) at day 7 vs. day 2 post-wounding in both PBO and 11β-HSD1 inhibitor AZD4017 (AZD) treated groups (PBO set 1: 99.7±0.8 vs. 56±27, set 2: 99.3±2.4 vs. 64.6±19.7, both p<0.001, and AZD set 1: 100±0 vs. 65.3±23.6, p<0.001, set 2: 100±0 vs. 84.7±13.5, all p<0.01, Figure 4b and 4d).

At 7 days post-wounding, re-epithelialization was complete in most cases and hence no conclusion on treatment effect could be made. However, in day 2 wounds after 30 days treatment, re-epithelialization was 30% greater in the AZD group (p<0.05).

#### Automated volumetric analysis

Representative histograms displaying wound sub-type area for each scan frame (120 frames per scan) by treatment and treatment day are presented in Figure 5.

**Fig. 5.**
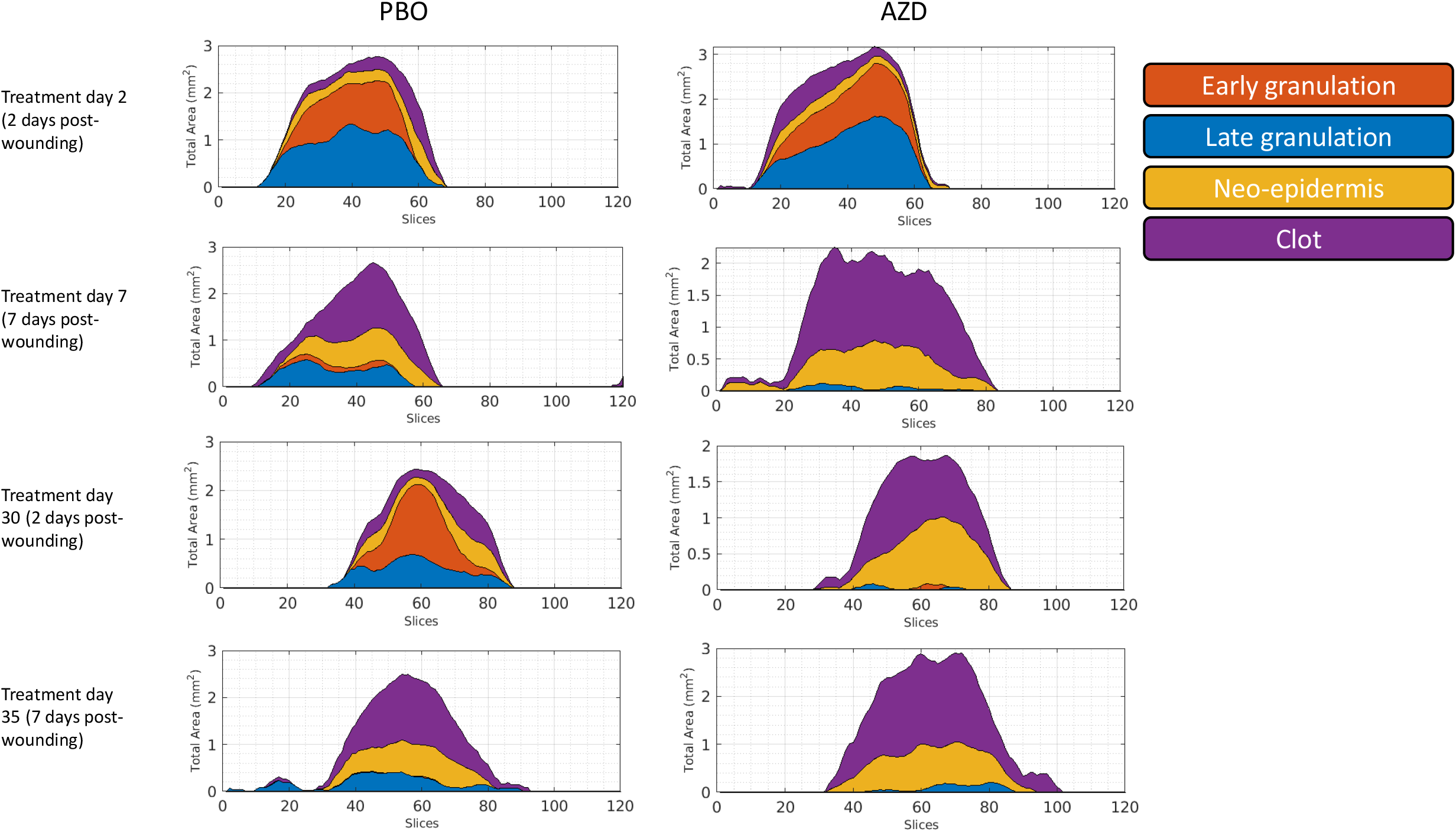
Machine learning histograms. Representative histograms for day 2 and day 7 post-wounding following 2-and 28-days’ treatment with AZD4017 (AZD) or placebo (PBO).

##### Early Granulation Tissue

Early granulation tissue volume (mean±SD) was significantly decreased at day 7 vs. day 2 post-wounding in both PBO and AZD treated groups and in both sets of biopsies (PBO set 1: 0.09±0.09 vs. 1.56±0.7, set 2: 0.13±0.17 vs. 1.38±0.7, both p<0.001 and AZD set 1: 0.1±0.1 vs. 1.36±0.7, p<0.001, set 2: 0.11±0.2 vs. 0.96±0.6, p<0.01, Figure 6a).

**Fig. 6.**
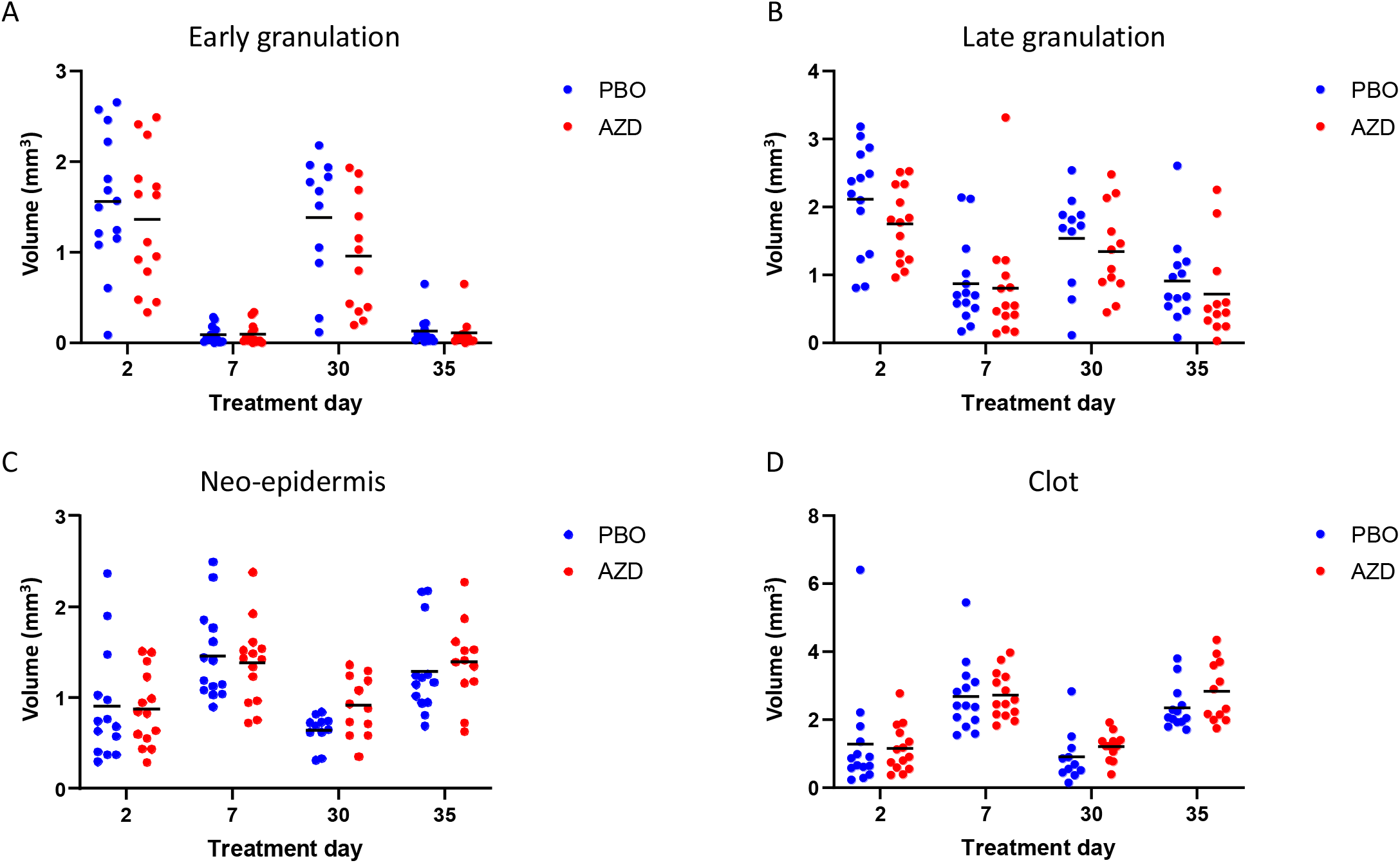
Machine learning volumetric quantification. Machine learning outputs for early (A) and late (B) granulation, neo-epidermis (C) and clot (D) tissue, comparing between day 2 (treatment day 2 and 30) and day 7 (treatment day 7 and 35) wounds treated with AZD4017 (AZD) or placebo (PCB).

At 7 days post-wounding there was no evidence of an AZD effect and resolution of early granulation tissue was mostly complete in all cases. In day 2 wounds after 30 days treatment, the effect with 11β-HSD1 inhibitor treatment was in the expected direction, but not statistically significant (p=0.48). This was also the case in day 2 wounds after 2 days treatment (p=0.93).

##### Late Granulation Tissue

A similar effect was observed for late granulation tissue volume (mean±SD), although volumes were higher overall, indicative of a more advanced stage of healing (as early granulation tissue is processed by resident fibroblasts to a more mature form). Day 7 vs. day 2 post-wounding late granulation tissue volume was PBO set 1: 0.87±0.6 vs. 2.12±0.8, p<0.001, set 2: 0.91±0.6 vs. 1.54±0.7, p=0.25 and AZD set 1: 0.81±0.8 vs. 1.75±0.5, p<0.05, set 2: 0.72±0.7 vs. 1.34±0.7, p<0.001 (Figure 6b).

At 2 days post-wounding and 2 days treatment, there was evidence of an effect with AZD4017 in the expected direction, but this was not statistically significant (p=0.53). No evidence of an effect was present at other time points (although variance was relatively high).

##### Neo-epidermis

In contrast to granulation tissue, neo-epidermis volume (mean±SD) was significantly increased at day 7 vs. day 2 post-wounding in all groups (PBO set 1: 1.46±0.5 vs. 0.9±0.6, p<0.01, set 2: 1.29±0.5 vs. 0.65±0.2, p<0.001 and AZD set 1: 1.38±0.4 vs. 0.87±0.4, p<0.01, set 2: 1.39±0.5 vs. 0.92±0.3, p<0.05, Figure 6c).

At 7 days post-wounding there was no evidence of an AZD effect and re-epithelialization was mostly complete in all cases. In day 2 wounds after 30 days treatment, there was a strong trend towards 42% greater neo-epidermal volume in the 11β-HSD1 inhibitor treatment group (p=0.09). This was not apparent in day 2 wounds after 2 days treatment (p=0.99).

##### Clot

Similarly to the neo-epidermis, clot volume (mean±SD) was found to significantly increase in day 7 vs. day 2 wounds, consistent with a more advanced stage of healing (PBO set 1: 2.68±1 vs. 1.28±1.6, set 2: 2.35±0.6 vs. 0.91±0.7, both p<0.001 and AZD set 1: 2.73±0.7 vs. 1.16±0.7, p<0.001, set 2: 2.83±0.9 vs. 1.21±0.4, p<0.01, Figure 6d).

In day 7 wounds after 35 days treatment, the change in clot volume with AZD4017 was in the expected direction, but not statistically significant (p=0.45). This was also true in day 7 wounds after 7 days treatment (p=0.99).

## Discussion

OCT has recently been validated as a non-invasive method of monitoring skin wound healing though a “virtual biopsy” (*29-33*) but volumetric analysis and automated classification wound healing had not previously been described. Here, we applied machine learning for automated quantification of key morphological features during wound repair.

Our findings suggest that OCT is a more sensitive method of monitoring wound healing, able to visualise key morphological features of wound healing otherwise undetectable using traditional imaging (e.g. digital photography). Despite a small sample size, we detected significant reductions in early clot / early granulation tissue and late granulation tissue with a concomitant increase in neo-epidermis and clot tissue volumes in day 7 wounds compared to day 2. These findings were highly reproducible between the two sets of biopsies in our cohort and importantly, reflect the normal healing chronology measured by two-dimensional OCT (*29*). However, our novel method of automated quantification has the key advantage of measuring the entire volume of specific wound healing features, which may not be accurately reflected by two-dimensional measurement.

Given the complex nature of the wound environments, inter-observer reliability (i.e. investigator agreement with machine annotation) was excellent for early granulation tissue, late granulation tissue and neo-epidermis, and good for clot (which exhibited a more heterogeneous morphology). Results were validated against manually derived measurements of early granulation tissue maximal width and maximal clot depth which were the only reportable healing outcomes of GC-SHEALD. The former correlated well with automated measures of volume but this was not evident for clot, likely due to the much narrower data range and lower machine learning sensitivity.

Punch biopsy healing in our T2DM cohort was largely normal, with complete re-epithelialization by day 7 in most samples and full healing (including blood clot detachment) by day 28. This was not unexpected, as patients with active ulceration and more poorly controlled diabetes (i.e. the intended target population) were excluded from GC-SHEALD pending safety results (which now support a future trial in patients with diabetic foot ulcers). However, the inclusion of patients with relatively normal healing may limit the scope of AZD4017 effectiveness. This is supported by our pre-clinical finding that topical 11β-HSD1 inhibition partially restores normal healing in mice treated with oral corticosterone (active rodent glucocorticoid) but has no benefit on normal mouse healing (despite comparable 11β-HSD1 inhibition)(*21*).

Despite this limitation, we found a 30% improvement in re-epithelialization by AZD4017, with a comparable trend in neo-epidermis volume. These additional analyses (including an automated and unbiased method) strengthen the core GC-SHEALD wound healing outcome that AZD4017 reduced wound gap width by 48% (*28*). Our results are also in agreement with mechanistic findings that glucocorticoids impair epidermal re-epithelialization through suppression of keratinocyte growth factor signalling (*15, 16*). However, the statistical interpretations of our results are based on a relatively small sample size and findings should be considered preliminary. Based on treatment day 30 results, estimated sample sizes of 43, 257, 12 and 37 per treatment group for early granulation, late granulation, neo-epidermis and clot (respectively) are anticipated to confirm the observed differences (90% power, alpha error 0.05).

In summary, our results demonstrate that automated volumetric quantification by OCT is a powerful technique, offering deeper insights into wound healing morphology. This now warrants further development in larger cohorts, with an extended acute wound healing time course in young healthy skin compared to complex wounds, such as diabetic foot ulcers. Through non-invasive monitoring of three-dimensional healing in real-time, clinicians could gain a more detailed understanding of treatment requirements, responses and disease-specific healing outcomes. Finally, our promising findings may lead to the discovery of novel prognostic biomarkers of healing for a more tailored approach to wound care management.

## Materials and Methods

### Study Participants

Data was acquired from the GC-SHEALD trial (www.isrctn.com/ISRCTN74621291 and https://clinicaltrials.gov/ct2/show/NCT03313297) with approval by the North West -Greater Manchester Central Research Ethics Committee (17/NW/0283) and following full informed consent. Briefly, participants were randomized in a double-blind manner to placebo (PBO, n=14) or 400mg bi-daily of the 11β-HSD1 inhibitor AZD4017 (AZD, n=14) for 35 days treatment. At days 0 and 28, wounds were induced by 2mm diameter full-thickness punch biopsy under local anaesthesia. Wounds were treated with a breathable dressing for 24 hours and imaged at 2-and 7-days post-wounding (treatment days 2, 7, 30 and 35).

### Optical Coherence Tomography

Scans were obtained using a Michelson-Diagnostics (Maidstone, UK) VivoSight™ scanner capturing 120 image frames 50µm apart (6mm). This was sufficient to capture each 2mm punch biopsy site in full. A total of 207 scans were available, taken from two biopsies at each time point per patient. Three scans were removed prior to machine learning input due to image acquisition anomalies e.g. both biopsies captured in one scan. Data outputs were screened for inter-observer reliability resulting in the exclusion of a further 17 scans, with the remaining scans representing 101/104 (>97%) averaged patient time points as follows: PBO treatment day 2 (n=14), PBO treatment day 7 (n=14), PBO treatment day 30 (n=11), PBO treatment day 35 (n=13), AZD treatment day 2 (n=14), AZD treatment day 7 (n=14), AZD treatment day 30 (n=12), PBO treatment day 35 (n=12).

Re-epithelialization data was based on surface OCT imaging data. Gross wound morphology was obtained by simultaneous aerial camera image capture during the OCT scan.

### Machine Learning

Within each scan frame, 7 distinctive image sub-types were defined: neo-epidermis, clot, early and late granulation tissue, intact tissue (peripheral to the wound), active bleeding and non-tissue. We used a u-net based convolutional neural network for the image segmentation of each 2D OCT frame, and the ground truth labels are annotations of the 7 sub-types (*34*). We acknowledge that other networks can also be used, including variations of the u-net (*35*) or the GANs model (*36*) and for simplicity in this first study of OCT data, we used classical u-net architecture.

U-net model training used 84 OCT frames annotated with the above 7 image sub-types, chosen manually from 207 scans (each scan comprising 120 frames). The training sample therefore represented 0.34% of available frames used in the study. The u-net architecture was written in Matlab 2019b, using a Linux cluster with three Tesla K80 GPUs training. This took 100 epochs with 1344 iterations per epoch, using a stochastic gradient descent with momentum=0.9, optimising at an initial learning rate of 0.05. The Factor for L2 regularization was set to 0.0001. The minimum batch size was set to 16 images and the training data was shuffled at every epoch. The training took 3483 minutes with a mini-bath accuracy of 85.85% at the final iteration.

The prediction of all samples was then performed using the trained model followed by a set of morphological operations to remove isolated pixels and small islands. Boundaries between classes were also smoothed with a Fourier descriptor. Considering the penetration depth of OCT scanner, we sampled to a depth of 1mm for each frame, calculating neo-epidermis, clot and granulation tissue area (mm^2^). Intact tissue, active bleeding (in a negligible number of samples) and non-tissue were excluded from the analysis. The areas were then multiplied by the scanning interval (50µm) for volumetric results (mm^3^).

### Statistical Methods

All data groups followed a normal distribution. Grouped analyses were performed (95% confidence interval) using a two-way analysis of variance mixed-effects model with *post hoc* testing corrected for multiple comparisons using Sidak’s test (GraphPad Prism, La Jolla, CA).

Correlations were analysed using Pearson’s correlation testing (95% confidence interval).

## Data Availability

The authors confirm that the data supporting the findings of this study are available within the article and its supplementary materials.

## Author contributions

Yinhai Wang: Conceptualization, Data curation, Investigation, Methodology, Project administration, Resources, Software, Supervision, Validation, Visualization, Writing – review & editing, Adrian Freeman: Conceptualization, Resources, Supervision, Writing -Review & Editing, Paul M Stewart: Funding acquisition, Ramzi Ajjan: Supervision, Writing -Review & Editing, and Ana Tiganescu: Conceptualization, Formal analysis, Funding acquisition, Investigation, Methodology, Project administration, Supervision, Validation, Visualization, Writing -Original Draft, Writing -Review & Editing.

## Data and materials availability

All data associated with this study are available in the main text or the supplementary materials.

## Trial registration

International Standard Randomised Controlled Trial Number 74621291 www.isrctn.com/ISRCTN74621291. ClinicalTrials.gov Identifier: NCT03313297 https://clinicaltrials.gov/ct2/show/NCT03313297.

## Supplementary Material

Representative wounds for treatment day 2 placebo (S1-S3) and AZD4017 (S4-S6), treatment day 7 placebo (S7-S9) and AZD4017 (S10-S12), treatment day 30 placebo (S13-S15) and AZD4017 (S16-S18) and treatment day 35 placebo (S19-S21) and AZD4017 (S22-S24).

## References and Notes

1. J. F. Guest, G. W. Fuller, P. Vowden, Cohort study evaluating the burden of wounds to the UK’s National Health Service in 2017/2018: update from 2012/2013. BMJ Open 10, e045253 (2020).

2. M. Olsson, K. Jarbrink, U. Divakar, R. Bajpai, Z. Upton, A. Schmidtchen, J. Car, The humanistic and economic burden of chronic wounds: A systematic review. Wound repair and regeneration : official publication of the Wound Healing Society [and] the European Tissue Repair Society 27, 114–125 (2019).

3. M. K. Ali, K. M. Bullard, J. B. Saaddine, C. C. Cowie, G. Imperatore, E. W. Gregg, Achievement of goals in U.S. diabetes care, 1999-2010. The New England journal of medicine 368, 1613–1624 (2013).

4. N. C. D. R. F. Collaboration, Worldwide trends in diabetes since 1980: a pooled analysis of 751 population-based studies with 4.4 million participants. Lancet 387, 1513–1530 (2016).

5. D. G. Armstrong, M. A. Swerdlow, A. A. Armstrong, M. S. Conte, W. V. Padula, S. A. Bus, Five year mortality and direct costs of care for people with diabetic foot complications are comparable to cancer. J Foot Ankle Res 13, 16 (2020).

6. J. M. Robbins, G. Strauss, D. Aron, J. Long, J. Kuba, Y. Kaplan, Mortality rates and diabetic foot ulcers: is it time to communicate mortality risk to patients with diabetic foot ulceration? J Am Podiatr Med Assoc 98, 489–493 (2008).

7. S. Pedras, R. Carvalho, M. G. Pereira, Predictors of quality of life in patients with diabetic foot ulcer: The role of anxiety, depression, and functionality. J Health Psychol 23, 1488–1498 (2018).

8. P. Cavanagh, C. Attinger, Z. Abbas, A. Bal, N. Rojas, Z. R. Xu, Cost of treating diabetic foot ulcers in five different countries. Diabetes Metab Res Rev 28 Suppl 1, 107–111 (2012).

9. N. Hex, C. Bartlett, D. Wright, M. Taylor, D. Varley, Estimating the current and future costs of Type 1 and Type 2 diabetes in the UK, including direct health costs and indirect societal and productivity costs. Diabetic medicine : a journal of the British Diabetic Association 29, 855–862 (2012).

10. A. American Diabetes, Economic costs of diabetes in the U.S. In 2007. Diabetes care 31, 596–615 (2008).

11. E. Harris, A. Tiganescu, S. Tubeuf, S. L. Mackie, The prediction and monitoring of toxicity associated with long-term systemic glucocorticoid therapy. Current rheumatology reports 17, 513 (2015).

12. A. C. DeVries, T. K. Craft, E. R. Glasper, G. N. Neigh, J. K. Alexander, 2006 Curt P. Richter award winner: Social influences on stress responses and health. Psychoneuroendocrinology 32, 587–603 (2007).

13. A. M. Mercado, D. A. Padgett, J. F. Sheridan, P. T. Marucha, Altered kinetics of IL-1 alpha, IL-1 beta, and KGF-1 gene expression in early wounds of restrained mice. Brain, behavior, and immunity 16, 150–162 (2002).

14. R. M. Gallucci, T. Sugawara, B. Yucesoy, K. Berryann, P. P. Simeonova, J. M. Matheson, M. I. Luster, Interleukin-6 treatment augments cutaneous wound healing in immunosuppressed mice. Journal of interferon & cytokine research : the official journal of the International Society for Interferon and Cytokine Research 21, 603–609 (2001).

15. B. Lee, C. Vouthounis, O. Stojadinovic, H. Brem, M. Im, M. Tomic-Canic, From an enhanceosome to a repressosome: molecular antagonism between glucocorticoids and EGF leads to inhibition of wound healing. Journal of molecular biology 345, 1083–1097 (2005).

16. A. Sanchis, L. Alba, V. Latorre, L. M. Sevilla, P. Perez, Keratinocyte-targeted overexpression of the glucocorticoid receptor delays cutaneous wound healing. PloS one 7, e29701 (2012).

17. M. S. Bitar, T. Farook, S. Wahid, I. M. Francis, Glucocorticoid-dependent impairment of wound healing in experimental diabetes: amelioration by adrenalectomy and RU 486. The Journal of surgical research 82, 234–243 (1999).

18. Y. Oishi, Z. W. Fu, Y. Ohnuki, H. Kato, T. Noguchi, Molecular basis of the alteration in skin collagen metabolism in response to in vivo dexamethasone treatment: effects on the synthesis of collagen type I and III, collagenase, and tissue inhibitors of metalloproteinases. The British journal of dermatology 147, 859–868 (2002).

19. A. Tiganescu, M. Hupe, Y. J. Jiang, A. Celli, Y. Uchida, T. M. Mauro, D. D. Bikle, P. M. Elias, W. M. Holleran, UVB induces epidermal 11beta-hydroxysteroid dehydrogenase type 1 activity in vivo. Experimental dermatology 24, 370–376 (2015).

20. A. Tiganescu, M. Hupe, Y. Uchida, T. Mauro, P. M. Elias, W. M. Holleran, Increased glucocorticoid activation during mouse skin wound healing. The Journal of endocrinology 221, 51–61 (2014).

21. A. Tiganescu, M. Hupe, Y. Uchida, T. Mauro, P. M. Elias, W. M. Holleran, Topical 11 beta-Hydroxysteroid Dehydrogenase Type 1 Inhibition Corrects Cutaneous Features of Systemic Glucocorticoid Excess in Female Mice. Endocrinology 159, 547–556 (2018).

22. A. Tiganescu, A. A. Tahrani, S. A. Morgan, M. Otranto, A. Desmouliere, L. Abrahams, Z. Hassan-Smith, E. A. Walker, E. H. Rabbitt, M. S. Cooper, K. Amrein, G. G. Lavery, P. M. Stewart, 11beta-Hydroxysteroid dehydrogenase blockade prevents age-induced skin structure and function defects. The Journal of clinical investigation 123, 3051–3060 (2013).

23. A. Tiganescu, E. A. Walker, R. S. Hardy, A. E. Mayes, P. M. Stewart, Localization, age-and site-dependent expression, and regulation of 11beta-hydroxysteroid dehydrogenase type 1 in skin. The Journal of investigative dermatology 131, 30–36 (2011).

24. S. A. Morgan, E. L. McCabe, L. L. Gathercole, Z. K. Hassan-Smith, D. P. Larner, I. J. Bujalska, P. M. Stewart, J. W. Tomlinson, G. G. Lavery, 11beta-HSD1 is the major regulator of the tissue-specific effects of circulating glucocorticoid excess. Proceedings of the National Academy of Sciences of the United States of America 111, E2482–2491 (2014).

25. C. B. Brazel, J. C. Simon, J. P. Tuckermann, A. Saalbach, Inhibition of 11beta-HSD1 Expression by Insulin in Skin: Impact for Diabetic Wound Healing. J Clin Med 9, (2020).

26. J. K. Youm, K. Park, Y. Uchida, A. Chan, T. M. Mauro, W. M. Holleran, P. M. Elias, Local blockade of glucocorticoid activation reverses stress- and glucocorticoid-induced delays in cutaneous wound healing. Wound repair and regeneration : official publication of the Wound Healing Society [and] the European Tissue Repair Society 21, 715–722 (2013).

27. M. Terao, H. Murota, A. Kimura, A. Kato, A. Ishikawa, K. Igawa, E. Miyoshi, I. Katayama, 11beta-Hydroxysteroid dehydrogenase-1 is a novel regulator of skin homeostasis and a candidate target for promoting tissue repair. PloS one 6, e25039 (2011).

28. R. Ajjan, E. M. Hensor, K. Shams, F. Del Galdo, A. Abbas, J. Woods, R. J. Fairclough, L. Webber, L. Pegg, A. Freeman, A. Morgan, P. M. Stewart, A. E. Taylor, W. Arlt, A. Tahrani, D. Russell, A. Tiganescu, A randomised controlled pilot trial of oral 11β-HSD1 inhibitor AZD4017 for wound healing in adults with type 2 diabetes mellitus. medRxiv, 2021.2003.2023.21254200 (2021).

29. N. S. Greaves, B. Benatar, S. Whiteside, T. Alonso-Rasgado, M. Baguneid, A. Bayat, Optical coherence tomography: a reliable alternative to invasive histological assessment of acute wound healing in human skin? The British journal of dermatology 170, 840–850 (2014).

30. J. Holmes, S. Schuh, F. L. Bowling, R. Mani, J. Welzel, Dynamic Optical Coherence Tomography Is a New Technique for Imaging Skin Around Lower Extremity Wounds. The international journal of lower extremity wounds 18, 65–74 (2019).

31. N. S. Greaves, S. A. Iqbal, T. Hodgkinson, J. Morris, B. Benatar, T. Alonso-Rasgado, M. Baguneid, A. Bayat, Skin substitute-assisted repair shows reduced dermal fibrosis in acute human wounds validated simultaneously by histology and optical coherence tomography. Wound repair and regeneration : official publication of the Wound Healing Society [and] the European Tissue Repair Society 23, 483–494 (2015).

32. G. D. Glinos, S. H. Verne, A. S. Aldahan, L. Liang, K. Nouri, S. Elliot, M. Glassberg, D. Cabrera DeBuc, T. Koru-Sengul, M. Tomic-Canic, I. Pastar, Optical coherence tomography for assessment of epithelialization in a human ex vivo wound model. Wound repair and regeneration : official publication of the Wound Healing Society [and] the European Tissue Repair Society 25, 1017–1026 (2017).

33. S. Ud-Din, P. Foden, K. Stocking, M. Mazhari, S. Al-Habba, M. Baguneid, D. McGeorge, A. Bayat, Objective assessment of dermal fibrosis in cutaneous scarring, using optical coherence tomography, high-frequency ultrasound and immunohistomorphometry of human skin. The British journal of dermatology 181, 722–732 (2019).

34. O. Ronneberger, P. Fischer, T. Brox. (Springer International Publishing, Cham, 2015), pp. 234–241.

35. E. Shelhamer, J. Long, T. Darrell, Fully Convolutional Networks for Semantic Segmentation. IEEE Trans Pattern Anal Mach Intell 39, 640–651 (2017).

36. S. Kazeminia, C. Baur, A. Kuijper, B. van Ginneken, N. Navab, S. Albarqouni, A. Mukhopadhyay, GANs for medical image analysis. Artificial Intelligence in Medicine 109, 101938 (2020).

